# Internet search patterns reveal clinical course of COVID-19 disease progression and pandemic spread across 32 countries

**DOI:** 10.1101/2020.05.01.20087858

**Authors:** Tina Lu, Ben Y. Reis

## Abstract

Effective public health response to novel pandemics relies on accurate and timely surveillance of pandemic spread, as well as characterization of the clinical course of the disease in affected individuals. We sought to determine whether Internet search patterns can be useful for tracking COVID-19 spread, and whether these data could also be useful in understanding the clinical progression of the disease in 32 countries across six continents. Temporal correlation analyses were conducted to characterize the relationships between a range of COVID-19 symptom-specific search terms and reported COVID-19 cases and deaths for each country during the period of January 1 through April 20, 2020. Increases in COVID-19 symptom-related searches preceded increases in reported COVID-19 cases and deaths by an average of 18.53 days (95% confidence interval 15.98 to 21.08) and 22.16 days (20.33 to 23.99), respectively. Cross-country ensemble averaging was used to derive average temporal profiles for each search term, which were combined to create a search-data-based view of the clinical course of disease progression. Internet search patterns revealed a clear temporal pattern of disease progression for COVID-19: Initial symptoms of fever, dry cough, sore throat and chills were followed by shortness of breath an average of 5.22 days (95% confidence interval 3.30 to 7.14) after initial symptom onset, matching the clinical course reported in the medical literature. This is the first study to show that Internet search data can be useful for characterizing the detailed clinical course of a disease. These data are available in real-time and at population scale, providing important benefits as a complementary resource for tracking the spread of pandemics, especially during the early stages before widespread laboratory testing is available.

---

Accurate real-time surveillance of local disease spread is essential for effective pandemic response, informing key public health measures such as social distancing and closures, as well as the allocation of scarce healthcare resources such as ventilators and hospital beds.^1 2^ During the current COVID-19 pandemic, surveillance has relied primarily on direct testing of individuals using a variety of *de novo* laboratory testing methods developed specifically for the emergent pandemic.^3^

While laboratory testing serves as an important primary gauge of epidemic spread, it is subject to a number of important limitations: 1. During the early stages of a novel pandemic, laboratory tests specific to the novel pathogen do not yet exist and therefore must be developed *de novo*, leading to significant delays before test-based surveillance can begin. 2. Once a test has been developed, scaling the manufacturing, distribution and test processing capacity takes a significant amount of time, resulting in limited testing capacity in many countries.^4^ It is therefore difficult to achieve population-scale surveillance with laboratory testing in the crucial early stages of an emergent pandemic. 3. Even after tests are widely available, operational delays in test administration and processing make laboratory testing a lagging indicator relative to disease onset.^5^ 4. Laboratory testing often requires individuals to leave home and congregate at testing centers, increasing exposure both for those being tested and for the health workers administering the tests. 5. Laboratory tests are typically invasive, involving blood draws or deep nasal swabs, which are uncomfortable for patients and increase the risk of infection to healthcare workers.^6^ 6. Laboratory testing can be expensive, especially when testing large numbers of people.^5^ 7. Certain types of laboratory tests may need to be repeated due to issues of reliability.^7, 8^

These limitations present an opportunity for exploring additional surveillance approaches^9^ that could be used as complementary information sources alongside laboratory testing, especially during the critical early stages of a pandemic. Aggregated de-identified Internet search patterns have been used to track a wide range of health phenomena, including influenza^10^, MERS^11^, measles^12^, abortion^13^ and immunization compliance,^14^ and are a potential complementary source of information for surveilling pandemic spread. Previous uses of these data have yielded valuable lessons in their appropriate use, including avoiding non-specific search terms, keeping models straightforward and transparent, and avoiding complex models that increase the risks of overfitting. When harnessed appropriately, Internet search patterns possess a number of strengths relative to laboratory testing: 1. Search data are collected continuously, and are ready to be used as soon as a new pandemic emerges. 2. Data are available at population-scale in areas with sufficient Internet access and usage levels.^15^ 3. Internet search data are available in near-real time, typically the same day. 4. There is no need for individuals to travel to testing locations; people can stay at home, avoiding increased exposure to themselves and to healthcare workers. 5. No physical intervention is required. 6. Internet search data are available for free.

## Characterizing Disease Progression

In addition to pandemic tracking, during the early stages of an emergent pandemic it is also important to characterize the clinical course of symptoms in affected individuals. At the individual patient level, knowing the expected clinical course of a disease allows doctors to understand where an individual patient is in their expected disease course, and what can be expected for that patient in the future. For example, in the case of COVID-19, it is important to know when dyspnea (shortness of breath) is expected to occur for patients, so that appropriate planning of clinical care and resources can take place.

A number of clinical case studies have analyzed the day-by-day clinical course for COVID-19 patients: A study of 138 patients in Wuhan, China found that common initial symptoms include fever and dry cough, followed by dyspnea a median of 5.0 days later.^16^ A meta-review found that the average length from illness onset to dyspnea was 4.99 days, based on evidence from 179 COVID-19 patients.^17^ A study in the US found that dyspnea typically set in between the fourth and eighth day following initial symptoms.^18^ A study of 41 patients in Wuhan, China found that the median duration from illness onset to dyspnoea was 8.0 days.^19^ The CDC’s *Interim Clinical Guidance for Management of Patients with Confirmed Coronavirus Disease (COVID-19)* states “Among patients who developed severe disease, the medium time to dyspnea ranged from 5 to 8 days”.^20^ These studies relied on a limited number of patients in hospital settings, and were published weeks and months after the initial spread of the pandemic.

Unlike the above hospital-based studies, Internet search data have the potential to provide information on disease progression across a much larger population of patients, not only those seen in hospital settings. Understanding the expected sequence of symptoms at a population level could also help public health officials track the different stages of the onset of the pandemic as it takes hold in a new location. It would therefore be beneficial for pandemic tracking and clinical planning if disease progression of novel pandemics could be studied in a timely manner and at population scales.

In this study, we conducted a systematic study across 32 countries on six continents to determine whether Internet search patterns can be used as a complementary data source for tracking COVID-19 spread, and whether these data could also be useful in understanding the clinical progression of the disease.

## Methods

### Data Acquisition

We selected a diverse set of 32 countries from six continents (Table 1), in which sufficient search data volumes were available for the search terms of interest. We obtained data on reported COVID-19 cases and deaths for each of these countries from a publicly available dataset maintained by the Center for Systems Science and Engineering at Johns Hopkins University. ^21^

**Table 1.** Countries included in the study, categorized by continent.

| **Continent** | **Countries** |
| --- | --- |
| Africa | Nigeria, South Africa, Zimbabwe |
| Asia | India, Iran, Malaysia, Saudi Arabia, Turkey |
| Oceania | Australia, Indonesia |
| North America | Canada, Guatemala, Mexico, United States |
| South America | Argentina, Brazil, Chile, Colombia, Uruguay, Venezuela |
| Europe | Belgium, France, Germany, Hungary, Ireland, Italy, Netherlands, Poland, Spain, Sweden, Switzerland, United Kingdom |

We collected daily relative search volume (RSV) data on a per-country basis for the period of January 1, 2020 through April 20, 2020, from Google Trends^22^ using the pytrends API^23^. Since Google has limited availability in China, we also accessed search trend data from the search engine Weibo.^24^ We accessed data for the following common symptoms of COVID-19: “fever”, “cough”, “dry cough”, “chills”, “sore throat”, “runny nose” and “shortness of breath”, as well as the more general terms “coronavirus”, “coronavirus symptoms” and “coronavirus test”. Each search term was searched as an “exact phrase”. We also considered other less common symptoms such as loss of smell and loss of taste, but the search volumes on those terms were too sparse for many countries. As daily search data are inherently noisy, all search data were smoothed with a 7-day moving average.^25^

For translations of the English search terms into local languages, we recruited native speakers to translate each term into the following languages: Arabic, Mandarin Chinese, Dutch, French, German, Italian, Persian, Polish, Portuguese and Spanish. These languages covered almost all of the countries included in the study. For languages in which we were unable to recruit native speakers for translations, we used Google Translate^26^ and confirmed that sufficient data were available for these translated search terms on Google Trends.^22^ A complete table of search terms and translations for each country is provided in Supplementary Table S-1.

### Data Analysis: Pandemic spread

We conducted temporal correlation studies to study the relationships between Internet search data and reported COVID-19 cases and deaths. For each country and search term, we calculated the Pearson correlation coefficient between the time series of search volumes for that search term and the time series of COVID-19 cases. We then shifted the search term data by a variable lag, and identified the lag that yielded the highest correlation. We computed the mean of these optimal lags for each search term across all countries. We then repeated these analyses substituting reported COVID-19 deaths for reported COVID-19 cases.

### Data Analysis: Clinical Course of Illness

We investigated whether Internet search data could be used to characterize the clinical course of COVID-19 symptoms over time. “Coronavirus symptoms” was selected as the index search term since it peaked first relative to the other search terms in 22 out of 32 countries. For each country, the date of peak search volume for the index search term was defined as the index date, and the dates for all other search data for that country were defined in relation to that date (Day 0, Day 1, Day 2, etc.). With the data from all countries aligned by the index date, cross-country ensemble average curves were calculated for each search term. (e.g. Day 1 values for “fever” searches in each of the 32 countries were averaged together to calculate the Day 1 value of the ensemble average “fever” curve. The same for Day 2, etc.) The ensemble average curves for all search terms were then overlaid on one plot, providing a search-data-based view of the clinical course of illness.

Since studies in the medical literature report the number of days between initial symptom onset and shortness of breath (dyspnea) ^16 17 18 19 20^, we examined a range of possible search-term-based definitions for initial symptom onset, based on various combinations of the earliest-peaking search terms “fever”, “cough”, “coronavirus symptoms” and “coronavirus test” (Table 3).

## Results

### Internet search volume and COVID-19 cases and deaths

We begin by examining a few example countries in which the pandemic peaked at different times. Figure 1 shows search volumes for the terms “fever” and “dry cough”, alongside reported COVID-19 cases and deaths for China, Iran, Italy, United States and India. Even though outbreaks occurred at different times in each country, the temporal relationships between the search terms and reported COVID-19 cases and deaths remained similar across countries.

**Figure 1.**
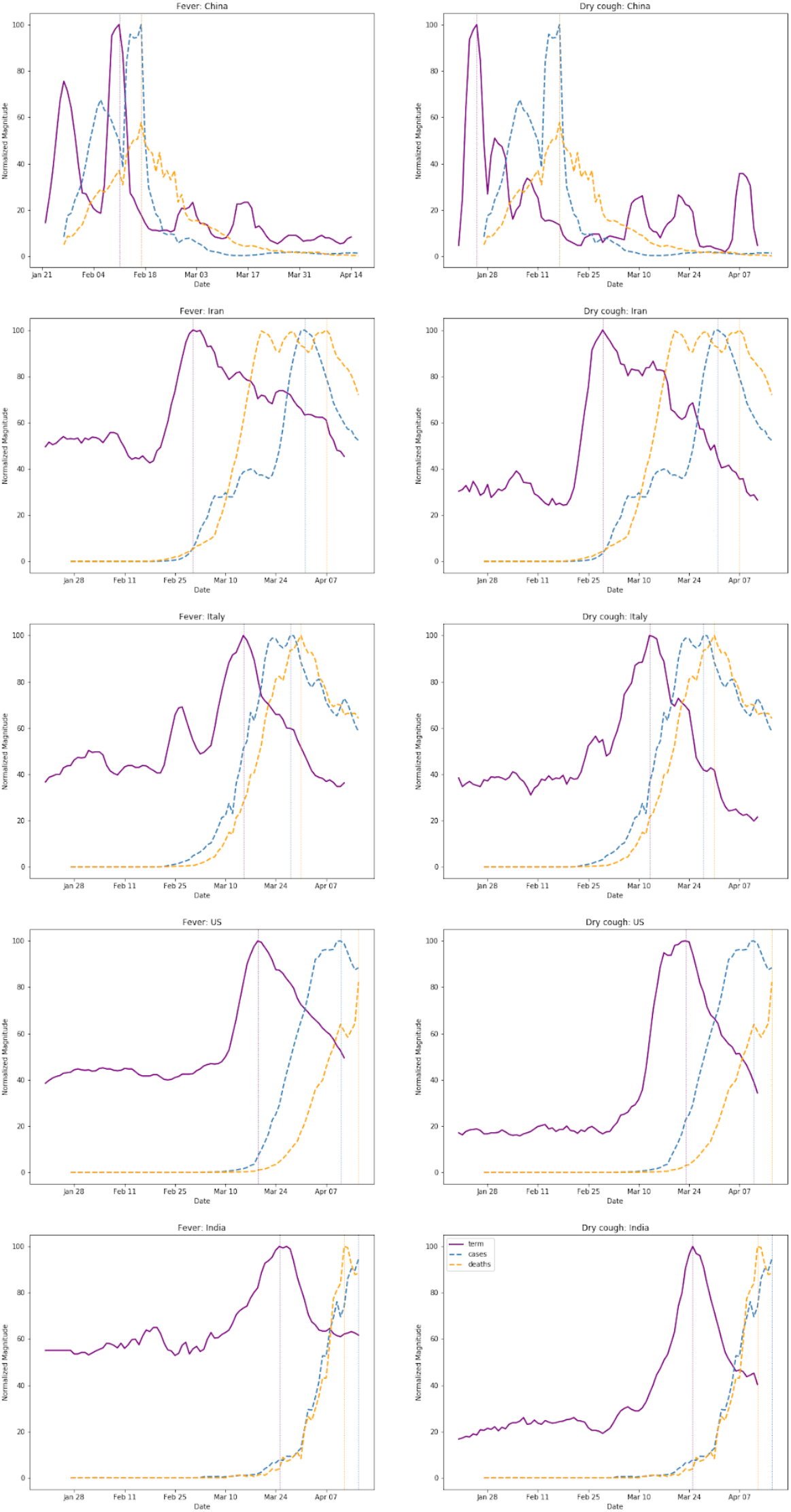
Search volumes (purple) for the terms “fever” (left) and “dry cough” (right), alongside reported COVID-19 cases (cyan) and deaths (orange) for China, Iran, Italy, US and India. Even though outbreaks occur at different times in different countries, the relationships between the search terms and reported COVID-19 cases and deaths remain similar across countries. To highlight the temporal relationships between the curves, the magnitude of each curve was independently normalized to fit the vertical dimensions of the plot.

Expanding this analysis to all 32 countries, Figure 2a shows the lags between search volumes for the term “fever” and COVID-19-related deaths for each of the 32 countries, along with a histogram showing the distribution of these lags. Figure 2b shows the lags between search volumes for the term “dry cough” and COVID-19-related deaths, along with the distribution of the lags.

**Figure 2.**
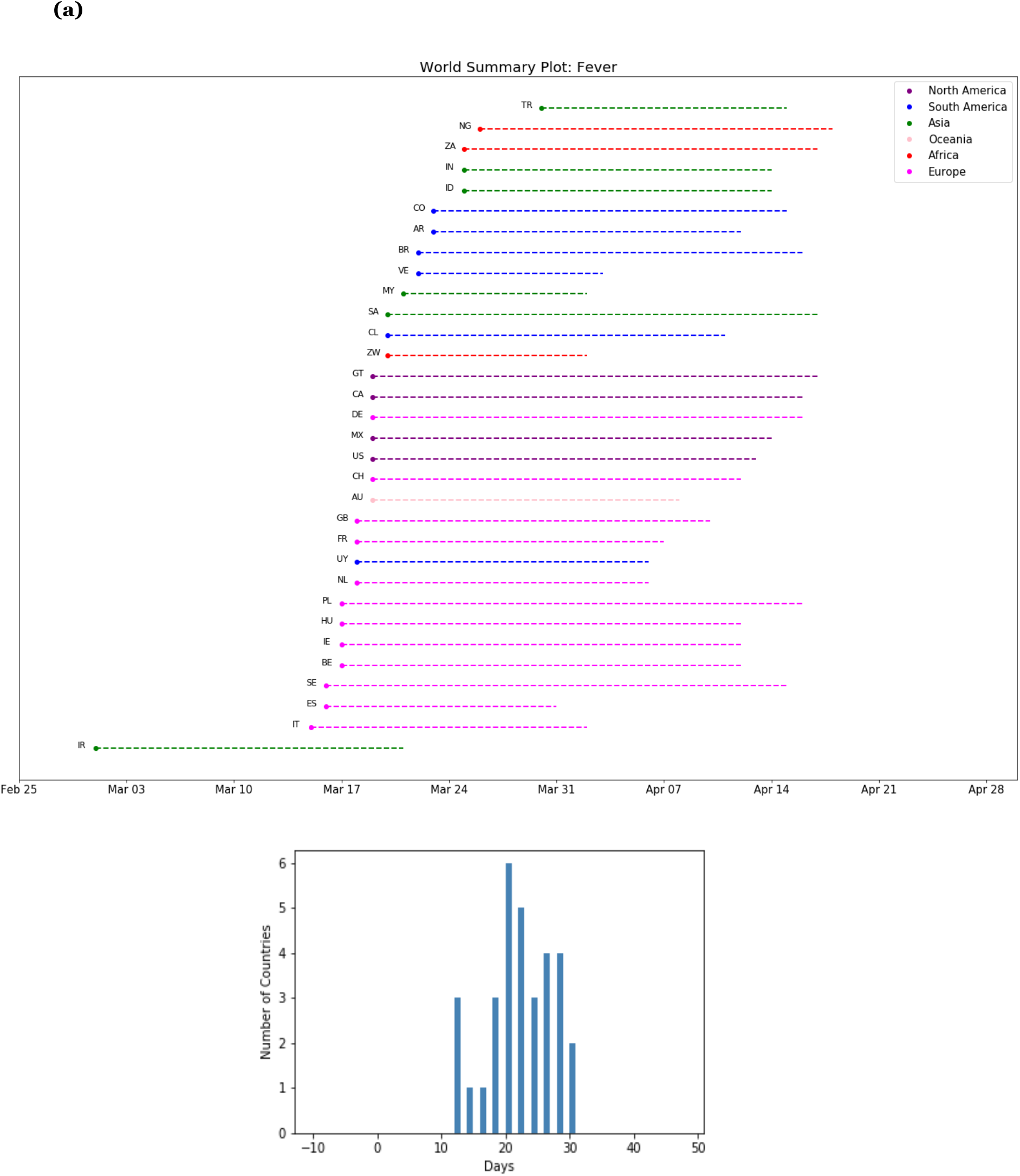
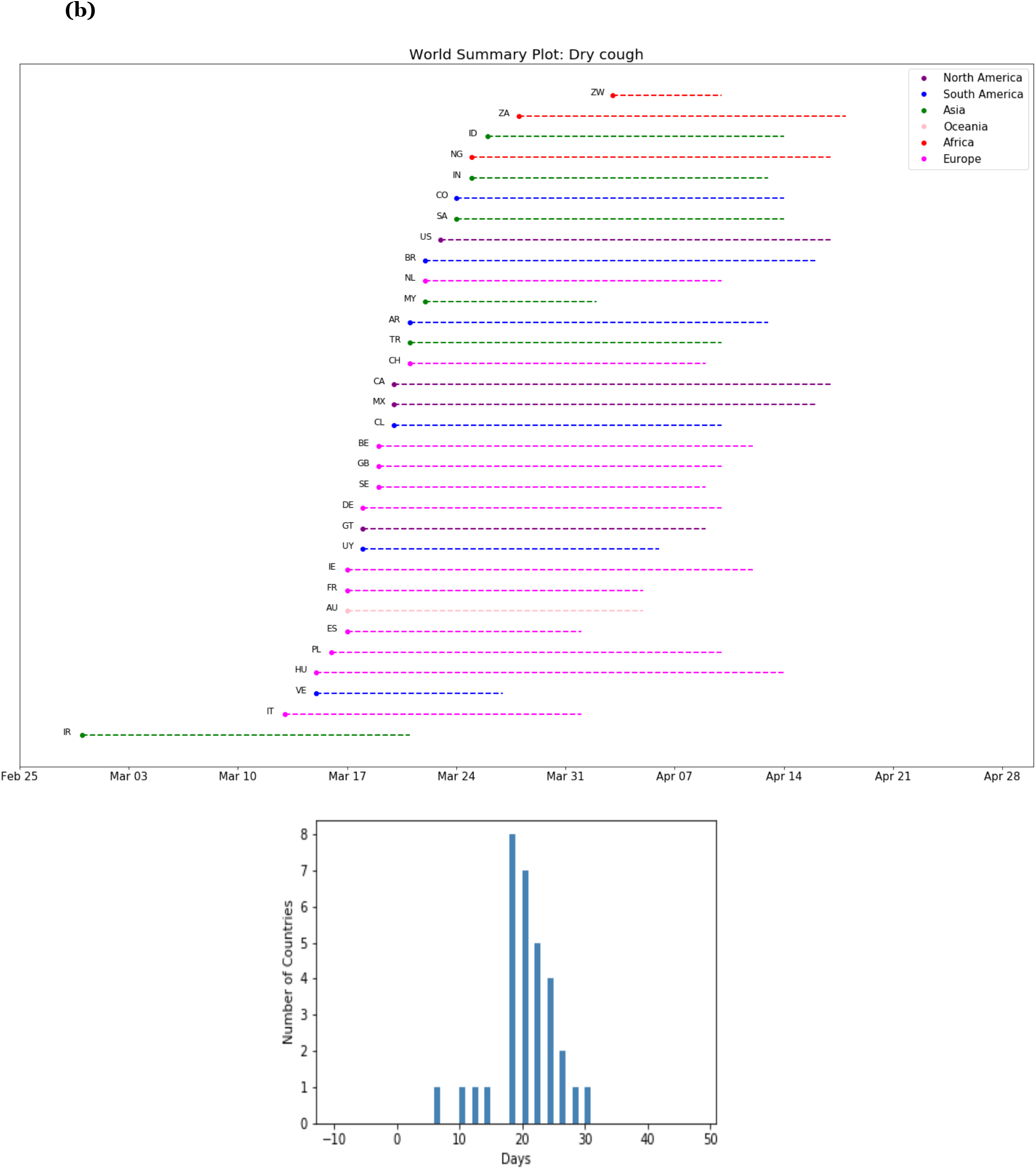
**(a)** Lags between searches for “fever” and reported COVID-19 deaths across 32 countries, with a histogram showing the distribution of these lags. Each country is labeled with its ISO Alpha-2 country code. **(b)** The same plots shown for searches for “dry cough”.

Table 2 shows summary statistics over all 32 countries for the lags between each of the ten search terms and COVID-19 cases and deaths, including mean and 95% confidence intervals. Searches for “fever” preceded increases in reported COVID-19 cases and deaths by an average of 18.53 days [95% CI 15.98-21.08] and 22.16 days [95% CI 20.33-23.99], respectively. For “cough” these times were 18.34 days [95% CI 16.05-20.64] and 21.88 days [95% CI 20.19-23.56], respectively. The complete list of lags between each search term and COVID-19 deaths for each individual country is provided in Supplementary Table S-2.

**Table 2.**
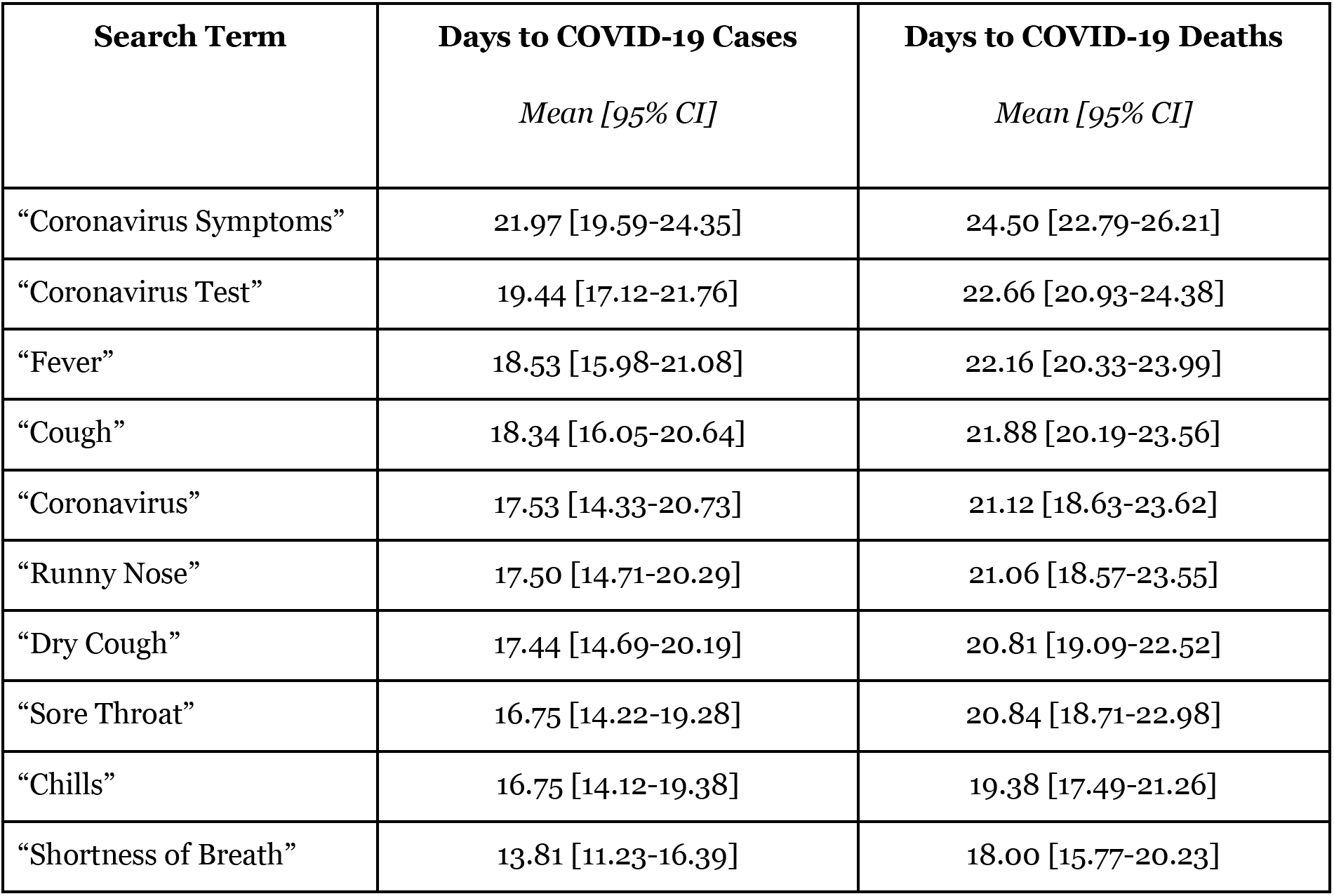
Mean lags between search volumes for specific search terms and reported COVID-19 cases and deaths across 32 countries. Mean lags are reported in days, along with 95% confidence intervals. Search terms are sorted chronologically, with those that peaked earliest appearing at the top of the table.

The average lags between searches and reported cases were shorter than those between searches and reported deaths, as cases are typically diagnosed and reported before deaths. We found that the inter-country variability of the average lags between searches and reported cases was greater (larger confidence intervals) than that for reported deaths, likely because case reporting is more dependent on local testing capacity and rates. We also found that the general term “coronavirus” has the greatest variability in its lags to reported cases and deaths (largest confidence intervals), compared to other symptom-specific terms, as would be expected for a more general search term.

### Internet Search Volume and COVID-19 Clinical Course of Illness

We begin by examining examples from individual countries, this time looking at the progression of symptom-related search terms over time. Figure 3 shows search volumes for a range of symptom-related search terms plotted over time for France (Figure 3a) and Mexico (Figure 3b). For each country, searches for “shortness of breath” appear a few days after searches for other symptoms such as “fever”, “cough” and “dry cough”.

**Figure 3.**
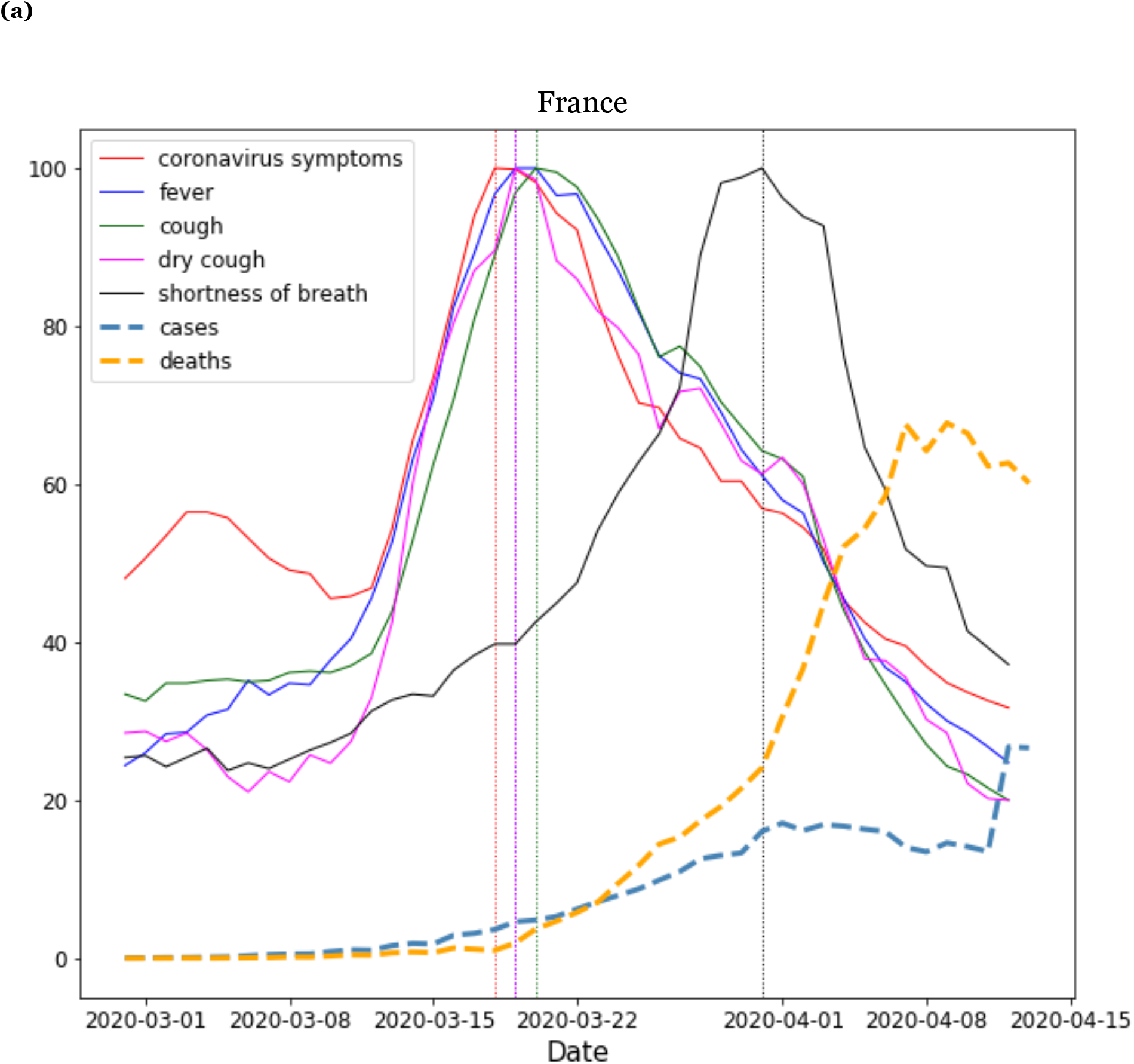
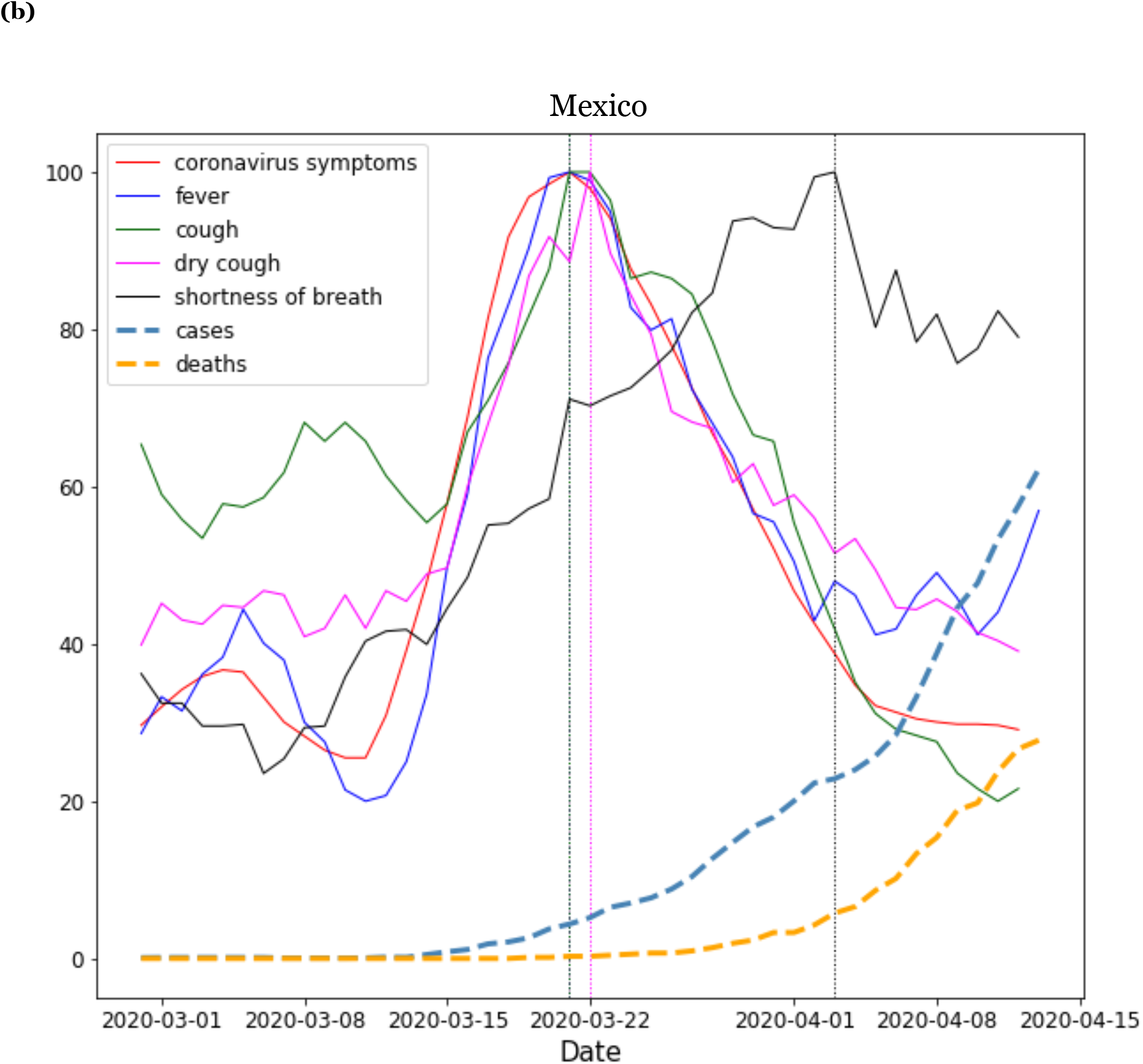
Clinical Course of COVID-19 symptoms in **(a)** France and **(b)** Mexico. Search volumes for the terms “coronavirus symptoms”, “fever”, “cough”, “dry cough” and “shortness of breath” (black) are shown alongside COVID-19 cases (dashed cyan line) and deaths (dashed orange line). Initial symptoms appear clustered together in time, with searches for shortness of breath appearing a few days later. To highlight the temporal relationships between curves, the magnitude of each curve was independently normalized to fit the vertical dimensions of the plot.

Expanding this analysis to all 32 countries, we calculated ensemble average curves for each search term across all 32 countries. Figure 4a shows the ensemble average search volumes for “fever”, “cough”, “dry cough” and “shortness of breath”, indexed by searches for “coronavirus symptoms”, alongside reported COVID-19 cases and deaths. Figure 4b shows this same analysis for additional search terms “sore throat”, “runny nose”, “chills”, and “coronavirus test”, also indexed by searches for “coronavirus symptoms”.

**Figure 4.**
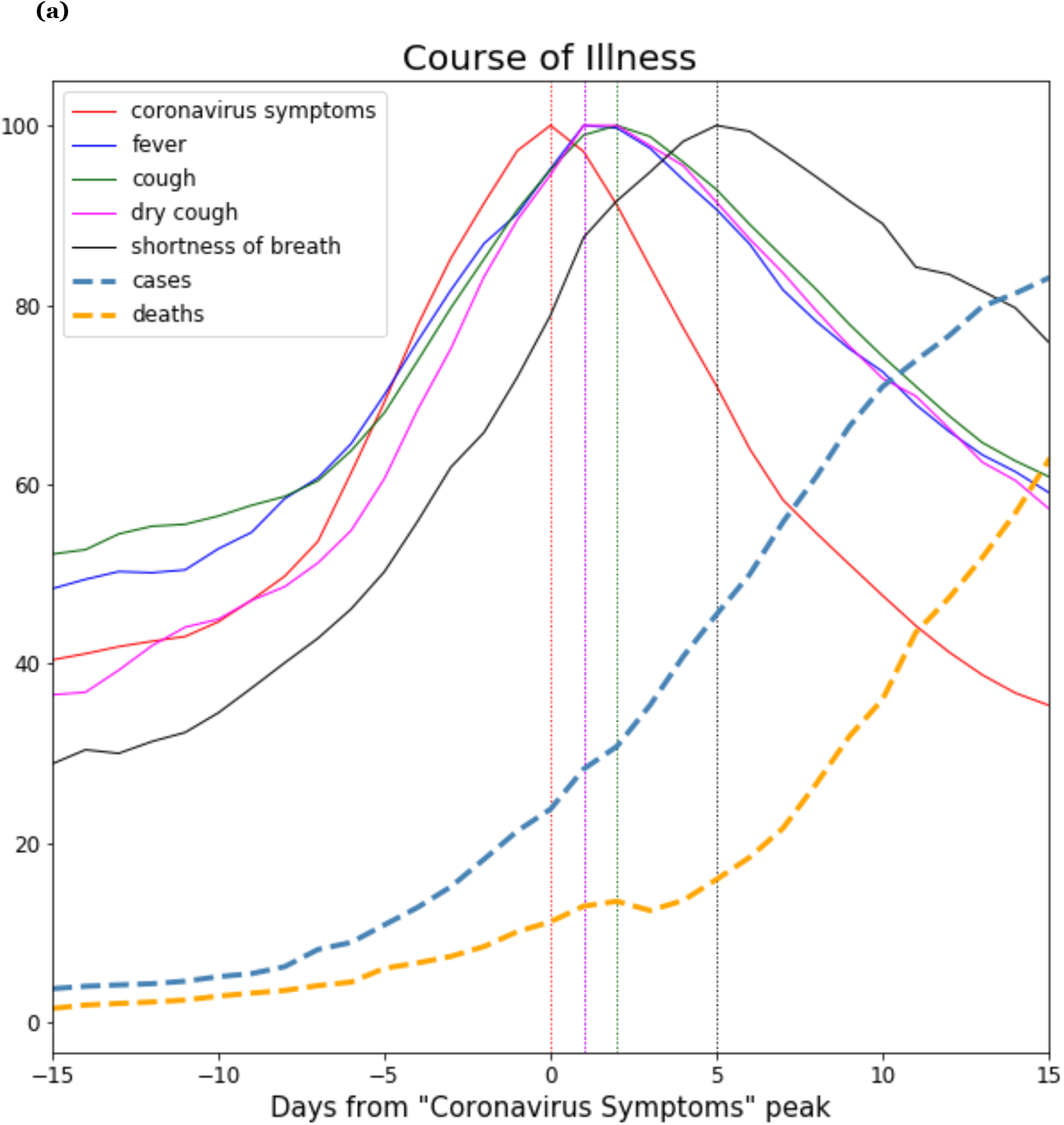
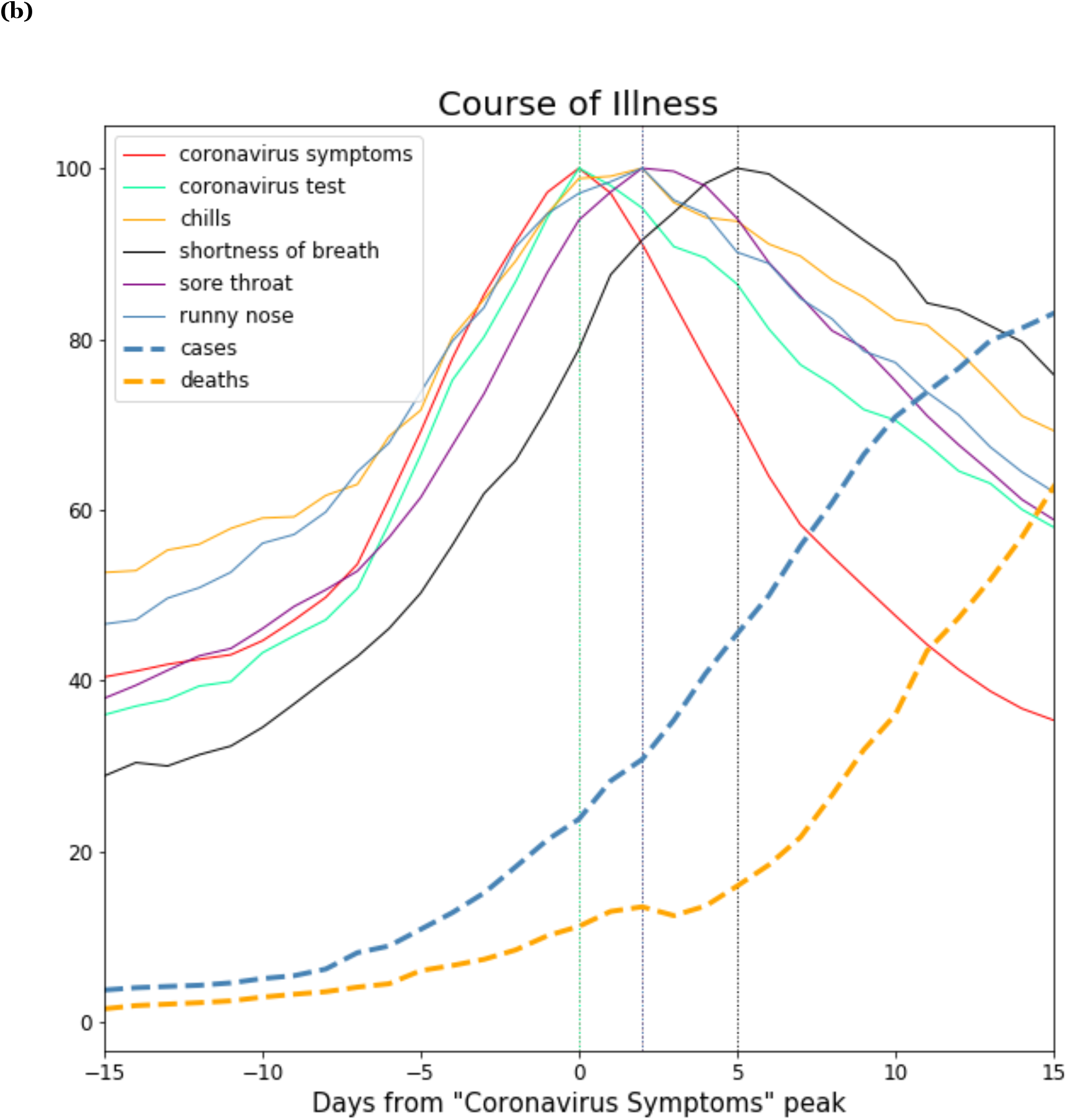
Ensemble average clinical course of COVID-19 symptoms across 32 countries, as seen through Internet search volumes of search terms related to COVID-19 symptoms. **(a)** Search volumes for the terms “fever”, “cough”, “dry cough”, “shortness of breath” (black), indexed by searches for “coronavirus symptoms”, shown alongside COVID-19 cases (dashed line cyan) and deaths (dashed line orange). **(b)** Search volumes for the terms “sore throat”, “runny nose”, “chills”, and “coronavirus test”, algonside “shortness of breath” (black), indexed by searches for “coronavirus symptoms”, shown alongside COVID-19 cases (dashed line cyan) and deaths (dashed line orange).

The clinical progression that emerges from these data presents the following picture: As the pandemic begins to take hold in a country, people search for “coronavirus symptoms” and “coronavirus test”, followed by initial symptoms “fever”, “cough”, “runny nose”, “sore throat” and “chills”, followed by searches for “shortness of breath” approximately 5 days after the search for initial symptoms.

Table 3 shows the lags between different definitions for initial symptom onset and searches for “shortness of breath”. The average lag between the searches for “fever” and “shortness of breath” was 5.22 days [95% CI 3.30-7.14]. For “cough” it was 5.16 days [95% CI 3.13-7.18]. These lags, as well as the lags deriving from other symptom onset definitions, are all around 5 days, matching the clinical course of the disease reported in the literature.^16 17 18 19 20^

**Table 3.** Different search-term-based definitions for symptom onset were examined by looking at different combinations of early-peaking search terms. The table shows average lag in days from various search-based definitions of symptom onset to searches for “Shortness of Breath” across 32 countries.

| **Symptom Onset**<br>*Search-Term-Based Definition* | **Days to “Shortness of Breath”**<br>*Mean [95% CI]* |
| --- | --- |
| “Fever” | 5.22 [3.30-7.14] |
| “Cough” | 5.16 [3.13-7.18] |
| Average of “Fever”, “Cough” | 5.19 [3.28-7.09] |
| Average of “Fever”, “Cough”, “Coronavirus Symptoms” | 5.81 [3.92-7.70] |
| Average of “Fever”, “Cough”, “Coronavirus Symptoms”, “Coronavirus Test” | 5.71 [3.85-7.57] |

The clinical progressions above are based on a 32-country ensemble average. While there is greater variability at the individual country level, the general temporal order of COVID-19 symptom progression remains fairly consistent across individual countries, with few exceptions. For example, only two of the 32 countries had searches for “coronavirus symptoms” peak after searches for “shortness of breath”, only 3 out of 32 countries had searches for “cough” peak after searches for “shortness of breath”, and only 4 out of 32 countries had searches for “fever” peak after searches for “shortness of breath” (Supplementary Table 2).

## Discussion

In this study, a systematic analysis of Internet search data from 32 countries across six continents found that increases in symptom-related Internet searches precede increases in reported COVID-19 cases and deaths by approximately 2-3 weeks. The present study also found that by analyzing data from all 32 countries, the overall temporal relationships between the different symptom-related search terms reflect the clinical progression of the symptoms of COVID-19.

To the best of the authors’ knowledge, the present study is the first to conduct a detailed investigation of multiple COVID-19 symptom-related search terms across such a large number of countries and languages. It is also the first to conduct a detailed analysis of the temporal relationships between different symptom-related searches in order to determine whether these data could be useful in understanding the clinical course of illness for a disease. During emergent pandemics, this detailed level of information can support health officials in tracking pandemic spread and planning clinical care and resources.

A number of recent studies have analyzed Internet search data related to the COVID-19 pandemic. Most of these studies examined data from a single country or from a small number of countries: Some studies examined only the search term “coronavirus” ^27, 28, 29 30^. In the present study we found that general non-symptom-specific search terms such as “coronavirus” have a greater variability in their relation to reported cases and deaths, likely due to the fact that individuals seeking general information on the pandemic may search for “coronavirus” even if they are not experiencing specific symptoms themselves at that time. Other studies looked at additional search terms such as “handwashing”, “face masks”,^31 32^ “quarantine”, “hand disinfection”,^33^ “SARS”, “MERS”, ^34^ “antiseptic”, and “sanitizer”^35^, but did not include specific symptom-related search terms.

Another group of studies did examine symptom-related search terms, including: “COVID pneumonia” and “COVID heart” in the US ^36^; “COVID” and “pneumonia” in China ^37^; “worry”, “hysteria” and other mental health symptoms in the US ^38^; “diarrhea”, “nausea”, “vomiting”, “abdominal pain”, “fever” and “cough” in the US^39^; “can’t smell” “ear pain”, “sinus pain”, “voice pain”, “ears ringing”. Among other terms in the US ^40^; “ageusia”, “abdominal pain”, “loss of appetite”, “anorexia”, “diarrhea”, and “vomiting” in the US^41^; “chest pain,” “myocardial infarction,” “cough,” and “fever” in four countries^42^; “Loss of sense of smell”,”Sense of smell”, “Loss of sense of taste”, and “Sense of taste” across multiple regions in five countries^25^; and “smell”, “loss of smell”, “anosmia”, “hyposmia”, “olfaction”, “taste”, “loss of taste”, and “dysgeusia” in eight countries ^43^. While these studies investigated a range of important questions, including the effects of COVID-19 on mental health^38^, gastrointestinal health ^39^,^41^, otolaryngological health^40^, and coronary-related conditions^42^, they did not study whether the temporal relationships between these search terms could be useful in understanding the clinical progression of the disease.

Some previous studies have tried to study temporal information from online data sources. For example, Liu et al. analyzed Twitter data to resolve different stages of behaviour relating to alcohol consumption (alcohol seeking, alcohol consuming, post-consumption reflection)^44^. Wu et al. studied seasonality in global public interest in psoriasis by analyzing seasonality in Internet search trends^45^. To the best of the authors’ knowledge, no previous studies have attempted to reconstruct the clinical progression of a disease based on Internet search data.

The use of Internet search data is subject to a number of important limitations.^46 10 47^ Internet infrastructure and digital access levels differ across countries and communities. Even though digital access rates are generally increasing worldwide, many developing countries currently lack sufficient search volumes to support search-based tracking. Search data may be subject to demographic, socio-economic, geographic, or other biases inherent in the local digital divide.^15^,^48^,^49^ In each country, the population of individuals who perform Internet searches may have different characteristics than those who do not, and the results inferred from Internet searching behaviors may not generalize to other populations.

Changes in search volumes for symptom-related terms such as “fever” could result not only from increases in COVID-19 cases, but also from general curiosity about the pandemic, the occurrence of other diseases (e.g. influenza, Lassa fever^50^), news coverage, or other factors. In this study, we used specific symptom-related search terms and examined data from a geographically diverse set of 32 counties across six continents and multiple languages. In recognition of the inherent variability of the data at the individual country level, we performed a global analysis combining data from all 32 countries. Despite the inherent variability of country-specific search data, our results show that the temporal relationships between the symptom-specific Internet search terms and COVID-19-related cases and deaths remained generally consistent between countries.

Future work includes applying these approaches to other diseases, and combining Internet search data with additional data sources such as local testing rates, public health measures, news reports, climatological and air quality variables, among others.

The ability of search data to not only help track pandemic spread, but also to reveal the clinical course of symptoms in emergent pandemics is significant. Given the limitations of laboratory testing, search data can be a valuable complementary source for population-scale tracking of pandemics in real time, particularly during the early stages of a pandemic when local testing is not yet available at scale, and can help to guide public health response.

## Data Availability

All data used in this study are publicly available through the sources referenced in the Methods section.

## Author Contributions

B.Y.R. and T.L conceived of the study, supervised its conduct, and oversaw data collection. B.Y.R. and T.L. designed the study, conducted statistical analysis, drafted the manuscript and formulated the implications of the results. All authors contributed substantially to the revision of the manuscript and approved the final manuscript as submitted.

## Competing Interest Statement

The authors declare no competing interests.

## Supplementary Materials

**Table S-1.**
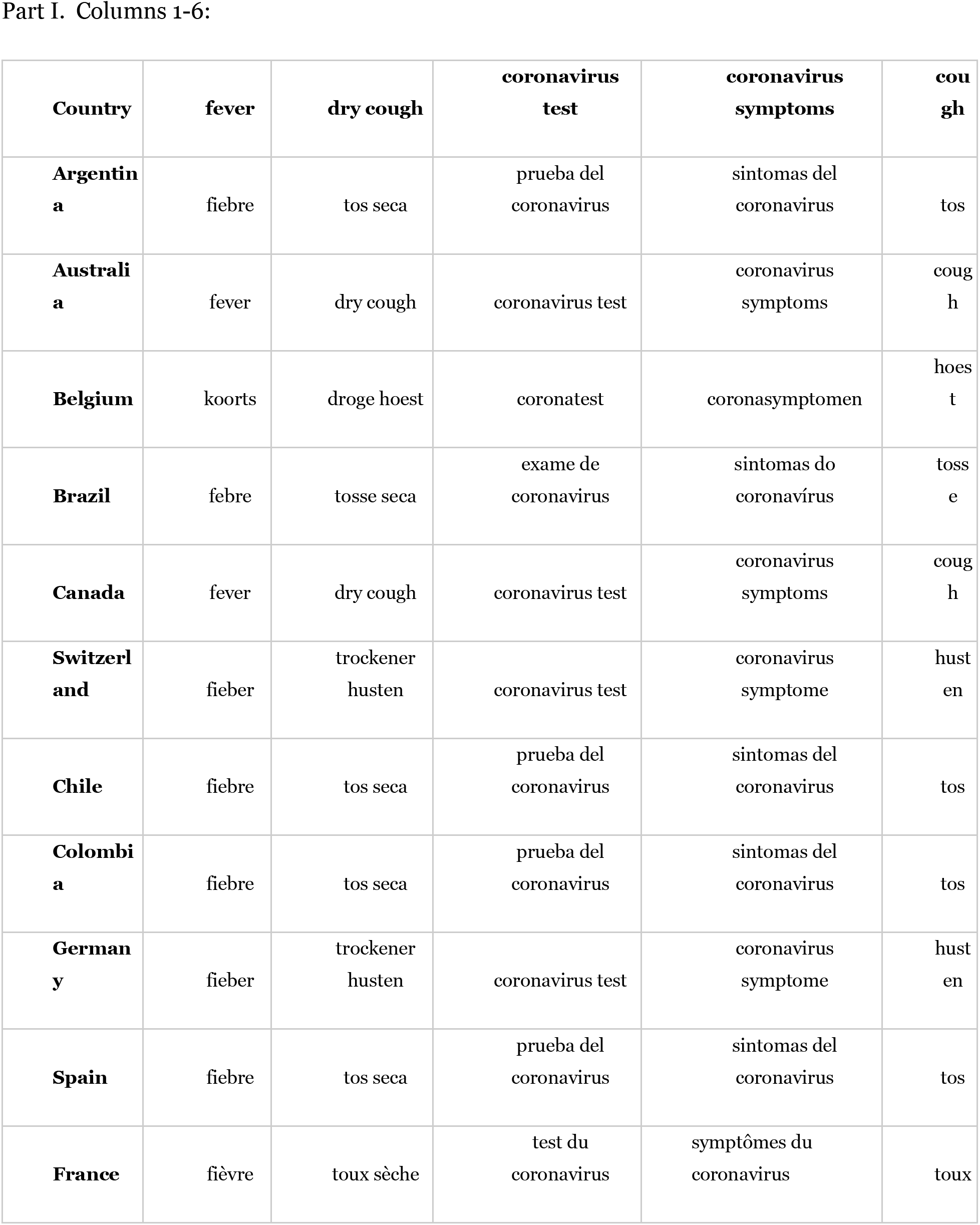
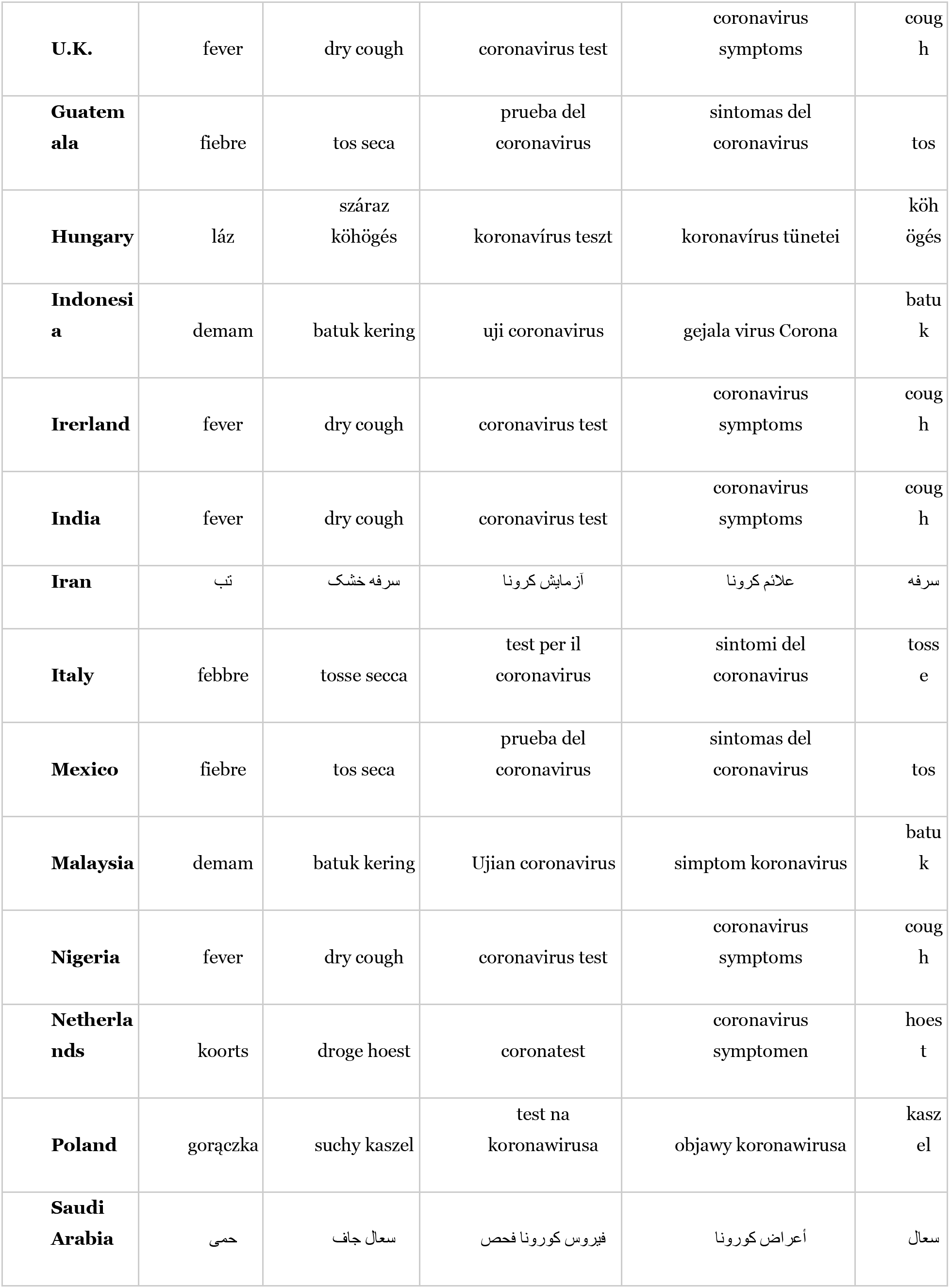
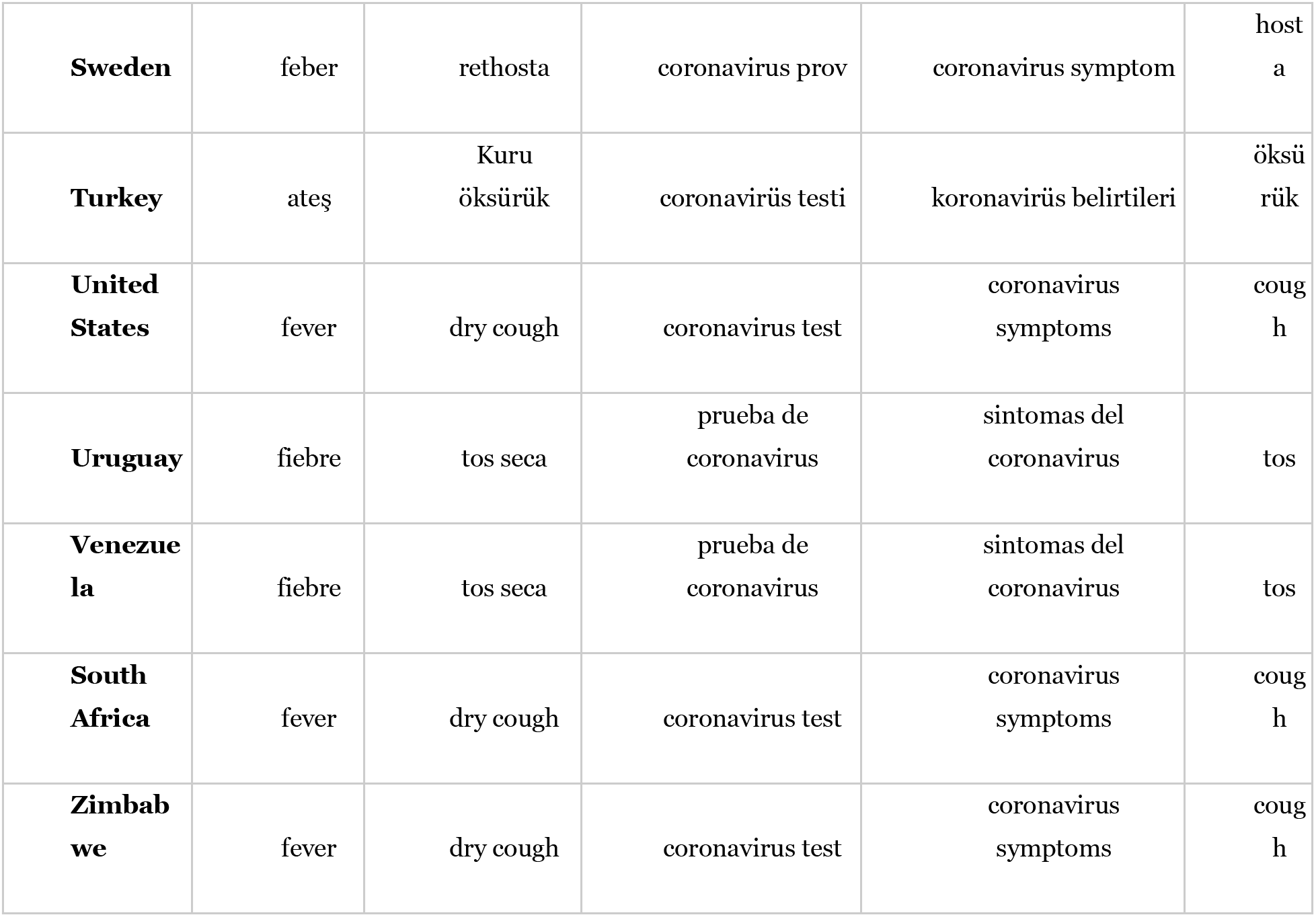
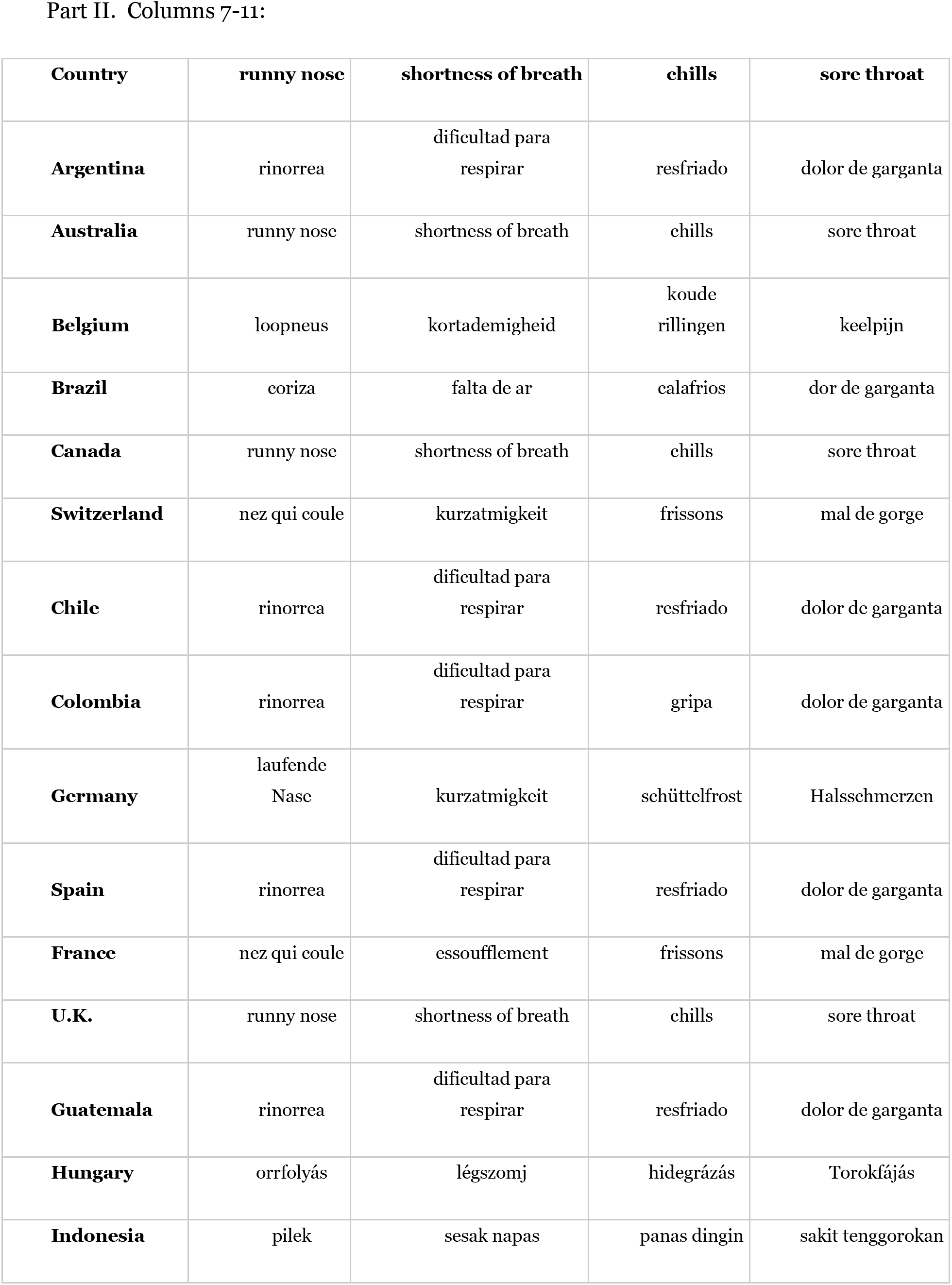
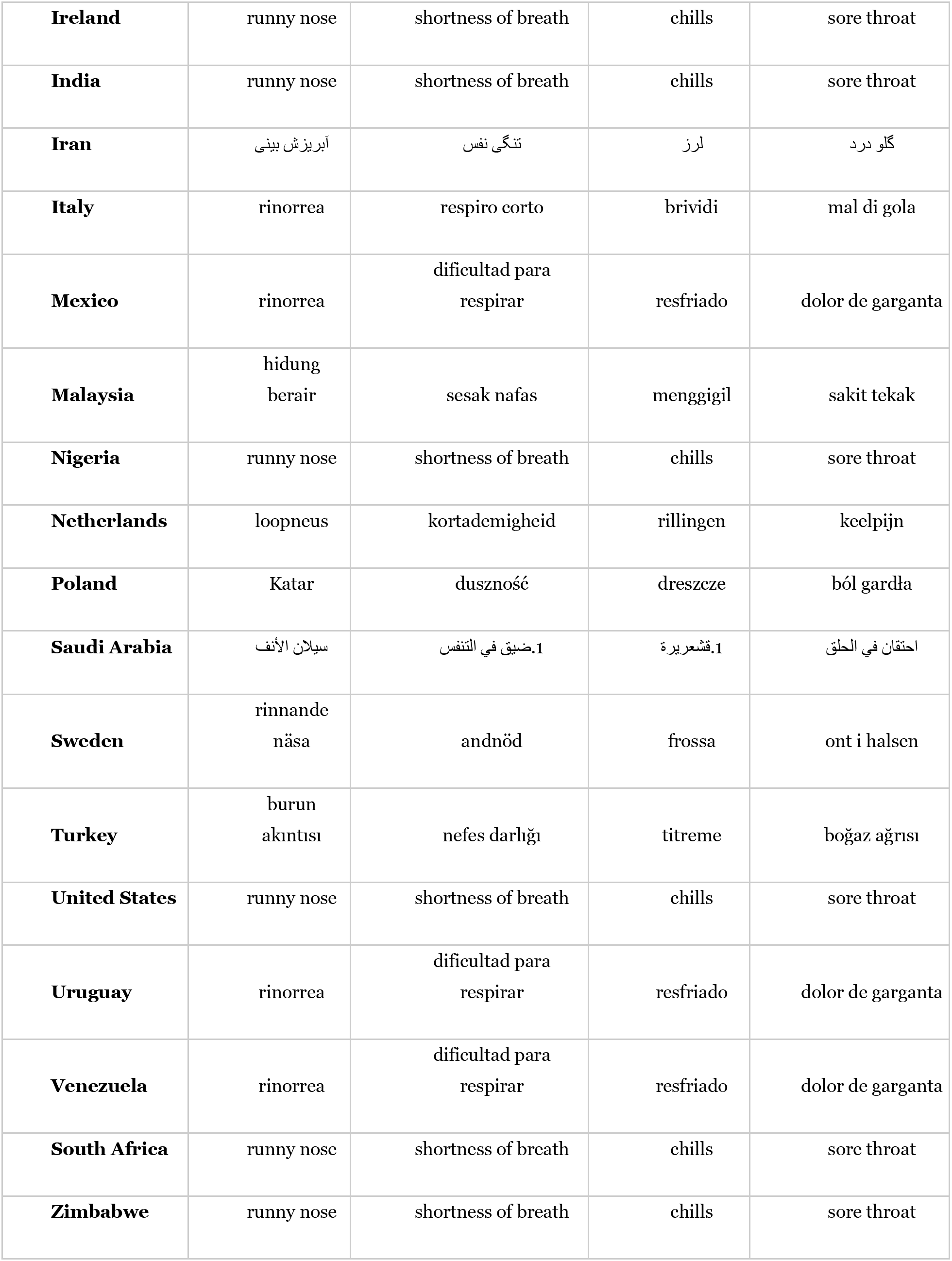
Translations used for each search term in each country.

**Table S-2.** Comprehensive list of lags between each of the search terms and COVID-19 deaths, for each of the 32 countries.

| Country | Coronavirus symptoms | Coronavirus test | Fever | Cough | Runny nose | Dry cough | Sore throat | Chills | Shortness of breath |
| --- | --- | --- | --- | --- | --- | --- | --- | --- | --- |
| **Argentina** | 24 | 19 | 20 | 21 | 24 | 23 | 22 | 26 | 21 |
| **Australia** | 23 | 22 | 20 | 18 | 20 | 19 | 19 | 16 | 20 |
| **Belgium** | 25 | 26 | 26 | 24 | 27 | 24 | 25 | 29 | 24 |
| **Brazil** | 21 | 28 | 25 | 25 | 25 | 25 | 25 | 22 | 18 |
| **Canada** | 30 | 30 | 28 | 27 | 29 | 28 | 28 | 22 | 27 |
| **Switzerland** | 25 | 20 | 24 | 25 | 17 | 19 | 23 | 13 | 15 |
| **Chile** | 24 | 21 | 22 | 22 | 23 | 21 | 23 | 24 | 20 |
| **Colombia** | 24 | 21 | 23 | 21 | 23 | 21 | 21 | 25 | 25 |
| **Germany** | 30 | 30 | 28 | 26 | 16 | 23 | 30 | 22 | 18 |
| **Spain** | 18 | 17 | 15 | 15 | 17 | 15 | 14 | 18 | 13 |
| **France** | 21 | 22 | 20 | 19 | 19 | 19 | 19 | 15 | 10 |
| **U.K.** | 27 | 23 | 23 | 22 | 23 | 22 | 22 | 22 | 22 |
| **Guatemala** | 29 | 22 | 29 | 30 | 25 | 22 | 26 | 29 | 13 |
| **Hungary** | 26 | 27 | 26 | 30 | 24 | 30 | 24 | 22 | 26 |
| **Indonesia** | 25 | 23 | 20 | 19 | 25 | 19 | 20 | 14 | 19 |
| **Ireland** | 30 | 28 | 26 | 25 | 29 | 26 | 25 | 25 | 25 |
| **India** | 28 | 24 | 20 | 19 | 25 | 19 | 27 | 17 | 17 |
| **Iran** | 25 | 22 | 20 | 21 | 28 | 21 | 20 | 16 | 17 |
| **Italy** | 23 | 27 | 18 | 24 | 8 | 19 | 20 | 12 | 11 |
| **Mexico** | 28 | 26 | 26 | 24 | 22 | 27 | 26 | 27 | 21 |
| **Malaysia** | 13 | 12 | 12 | 12 | 0 | 11 | 6 | 20 | 4 |
| **Nigeria** | 25 | 21 | 23 | 22 | 19 | 23 | 20 | 21 | 17 |
| **Netherlands** | 30 | 16 | 19 | 20 | 19 | 19 | 20 | 17 | 17 |
| **Poland** | 27 | 25 | 30 | 26 | 30 | 25 | 25 | 22 | 18 |
| **Saudi Arabia** | 23 | 23 | 28 | 23 | 14 | 21 | 3 | 20 | 10 |
| **Sweden** | 30 | 23 | 30 | 24 | 21 | 21 | 25 | 12 | 22 |
| **Turkey** | 21 | 21 | 16 | 19 | 23 | 20 | 18 | 14 | 18 |
| **United States** | 30 | 28 | 25 | 25 | 27 | 25 | 24 | 22 | 25 |
| **Uruguay** | 18 | 18 | 19 | 16 | 18 | 19 | 17 | 18 | 7 |
| **Venezuela** | 16 | 15 | 12 | 15 | 18 | 12 | 16 | 9 | 8 |
| **South Africa** | 30 | 30 | 23 | 29 | 29 | 21 | 22 | 13 | 18 |
| **Zimbabwe** | 15 | 13 | 13 | 12 | 5 | 7 | 12 | 11 | 30 |

## Notes

### Competing Interest Statement

The authors have declared no competing interest.

### Funding Statement

This research was made possible by funding from NIH grants R01HG009129, R01MH117599 and R01MH116042.

### Author Declarations

Only aggregate anonymized public Internet search trend data analyzed.

### Summary of Updates

Supplementary tables added with further details of analysis.

